# Efficacy of the Serogroup B *Neisseria meningitidis* Vaccine (4CMenB) in Preventing Experimental *Neisseria gonorrhoeae* Urethral Infection: A Double-Blind Randomized Controlled Human Challenge Study Protocol

**DOI:** 10.64898/2025.12.12.25341919

**Authors:** Andreea Waltmann, Catherine Kronk, Dana Lapple, Michael P. Motley, Feng-Chang Lin, Arlene C. Seña, Marcia M. Hobbs, Joseph A. Duncan

## Abstract

**Introduction:** Rates of sexually transmitted infections are on the rise globally, including those caused by *Neisseria gonorrhoeae*. Retrospective analyses across clinical settings around the world have indicated that rates of gonorrhea declined following mass vaccination campaigns vaccines made with outer membrane vesicles (OMV) from *Neisseria meningitidis* serogroup B, a pathogen closely related to *N. gonorrhoeae.* The US FDA-approved *N. meningitidis* serogroup B vaccine (4CMenB) contains *N. meningitidis* OMV and five recombinant protein components: fHbp (*factor H–binding protein*), NHBA (*Neisserial heparin-binding antigen*), NadA (*Neisseria adhesin A*), and the fusion partners GNA2091 and GNA1030 (genome-derived neisserial antigens used to stabilize fHbp and NHBA, respectively); *N. gonorrhoeae* encodes homologs of fHbp, NHBA, GNA2091, and GNA1030, but not NadA. Humans and mice immunized with 4CMenB have been shown to develop antibodies directed against *N. gonorrhoeae* antigens, and 4CMenB immunized mice exhibit enhanced *N. gonorrhoeae* clearance in a vaginal-infection model *vs*. mock immunized mice. Collectively, these data led to the hypothesis that immunization with 4CMenB may provide cross-protection against *N. gonorrhoeae.* There is a unique and time-sensitive opportunity to test the effectiveness of this vaccine in preventing *N. gonorrhoeae* using the human male urethral challenge model.

**Methods:** This is a double-blind randomized controlled trial testing the efficacy of the FDA-approved 4CMenB vaccine in preventing experimental *N. gonorrhoeae* infection. Our research team is currently the only in the world to utilize the established controlled human infection model to study *N. gonorrhoeae* in its natural human host. Males aged ≥18 and <36 years with no medical contraindication to 4CMenB administration, no contraindication to intraurethral challenge with *N. gonorrhoeae*, and who are willing and able to consent and participate in the study will be enrolled in this study. Participants who complete both immunization and *N. gonorrhoeae* challenge phases will be included in the primary endpoint analyses of *N. gonorrhoeae* infection in each study group to determine the efficacy of standard immunization with 4CMenB in preventing experimental gonococcal infection. The secondary objective is to determine whether 4CMenB vaccinations change or delay the onset of urethritis symptoms.

**Ethics:** All participants enrolled in the study provided informed consent twice. This study has been reviewed and approved by the University of North Carolina at Chapel Hill Institutional review Board (#21-0498). Data collection for this study is ongoing.

**Trial registration:** ClinicalTrials.gov Identifier: NCT05294588

**Article summary:** The paper presents the protocol for a double-blind randomized controlled human challenge study designed to evaluate the efficacy of the FDA-approved Serogroup B *Neisseria meningitidis* vaccine (4CMenB) in preventing experimental *Neisseria gonorrhoeae* urethral infection in men. This research addresses the urgent global public health threat posed by rising gonorrhea incidence and accelerating antimicrobial resistance (AMR), which highlights the critical need for vaccines. The study is motivated by retrospective analyses from clinical settings globally, which indicated that mass vaccination campaigns using *N. meningitidis* outer membrane vesicle (OMV) vaccines, including 4CMenB, correlated with a decline in symptomatic gonorrhea rates, with an estimated protective effect in the range of 30–40%. This observed cross-protection is biologically plausible because *N. gonorrhoeae* encodes homologs of several key protein components contained in the 4CMenB vaccine. The primary objective of the randomized trial is to formally determine the efficacy of standard 4CMenB immunization in preventing the acquisition of *N. gonorrhoeae* infection, defined by positive NAAT or culture. A secondary objective is to determine if 4CMenB immunization changes or delays the onset of urethritis symptoms. The study enrolls men aged 18 to 36 years who are randomized to receive two doses of either 4CMenB (experimental arm) or two doses of irrelevant FDA-approved control vaccines (quadrivalent influenza and tetanus/diphtheria). The investigation utilizes the controlled human infection model (CHIM) of gonorrhea, which is uniquely employed by the research team. This approach is justified by its safety record (over 30 years of safe use at UNC) and its superior statistical power compared to observational studies, allowing the trial to detect moderate vaccine efficacy with a much smaller number of participants. This CHIM design will not only deliver a direct efficacy estimate but also generate high-value mechanistic data by permitting detailed investigation of early infection events and immune correlates of protection in the natural human host.

## Introduction

*Neisseria gonorrhoeae* poses a synergistic global public health threat because of rising incidence and emerging antimicrobial resistance (AMR). *N. gonorrhoeae* is one of the most common sexually transmitted bacterial pathogens worldwide with roughly 90 million new cases occurring each year [1]. *N. gonorrhoeae* generally infects the urethra in men and cervix in women, but it can ascend into the upper reproductive tract including the epididymis and fallopian tubes. Control of *N. gonorrhoeae* infection (*i.e.,* gonorrhea) has relied on single-dose, directly observed antibiotic administration. However, AMR in *N. gonorrhoeae* remains widespread, including to ceftriaxone, the only remaining globally recommended first-line treatment for gonorrhea. Alarmingly, detection of extensively drug-resistant strains (to ceftriaxone and other antibiotic classes) is not only occurring but accelerating, with confirmed cases now reported in Australia, across Europe, the United Kingdom, and Asia [2]. Therefore, additional measures to control of this pathogen are urgently needed to reduce the public health burden of gonorrhea. Vaccines or immunotherapeutics may be the only sustainable solution into the future, given AMR has emerged in *N. gonorrhoeae* to all antibiotics ever deployed to treat its infection [3].

Effective vaccines for the most prevalent serogroups of *Neisseria meningitidis* have been in use in humans for decades. Meningococcal vaccines were first generated against polysaccharide capsular antigens of *N. meningitidis* serogroups A,C,Y, and W. *N. meningitidis* serogroup B (NmB) capsule is a poorly immunogenic disaccharide in humans, which limited development of vaccines targeting NmB [4]. In the past, vaccines containing several proteins and lipoproteins have been generated from NmB outer membrane vesicles (OMV) [5, 6]. These vaccines are safe and have been used in mass vaccination campaigns in the setting of NmB epidemics in several countries[7–9]. STI surveillance in the setting of Nmb-OMV vaccination in New Zealand, Cuba, Norway, and Australia, showed subsequent reduced rates for symptomatic gonorrhea [10–17]. This suggests that *N. meningitidis* OMV-based vaccines may offer cross-species protection against *N. gonorrhoeae*.

Currently, the *N. meningitidis* OMV-containing vaccine, 4CMenB (tradename Bexsero), is US FDA approved for the prevention of invasive MenB infection. Similar to observations made after *N. meningitidis* OMV only vaccines, recent retrospective studies that evaluated the effectiveness of 4CMenB against gonorrhea in Canada, Australia, and the United States have reported declines in gonorrhea incidence following vaccination [13, 18, 19]. Overall, the estimated protective effect of NmB-OMV vaccines, including 4CMenB, against symptomatic *N. gonorrhoeae* infection is estimated to be in the range of 30-40% [10–17]. A recent modeling study investigating the health and economic benefits of using 4CMenB against gonorrhea has found that even a moderately effective vaccine can have a significant public health impact [20]. However, it must be noted that these retrospective studies did not include active surveillance for gonorrhea and could be confounded by the presence of asymptomatic infections in the vaccinated populations. Thus, it is unclear whether the currently available data from retrospective studies reflect true clinical protection from all infections or are limited to persons with symptomatic gonorrhea. This is important to clarify because asymptomatic infections, which can comprise up to 45% of all infections, can have significant impacts on the individual (untreated infections can lead to serious sequalae), on public health (due to continued transmission), and on the economy (due to losses in productivity and costs to the healthcare system[21]).

A strong biological plausibility of cross-species protection exists. There is a high degree of genetic identity between meningococcal vaccine components and their gonococcal homologs. The OMV surface proteins of the meningococcal strain utilized as the OMV source in 4CMenB (NZ98/254) share a mean sequence conservation of 91.0% with their *N. gonorrhoeae* homologs [22]. In addition, 4CMenB contains three recombinant *N. meningitidis* protein antigens[22]: Neisseria Heparin Binding Antigen (NHBA) fused to genome-derived *Neisseria* antigen (GNA) 1030, Factor H Binding Protein (fHbp) fused to GNA 2091, and *Neisseria* adhesion A (NadA). NHBA, fHbp, and NadA are believed to be antigen targets of bactericidal antibodies responsible for protection from invasive *N. meningitidis* diseases [23–25]. NadA lacks a homolog in *N. gonorrhoeae*, but NHBA and fHbp in the vaccine have high genetic identity to *N. gonorrhoeae* homologs [22]. NHBA is known to be surface-expressed in *N. gonorrhoeae*[26]. While fHbp is expressed on the surface of *N. meningitidis,* the *N. gonorrhoeae* homolog appears to be an intracellular protein [27]. Vaccinating with fHbp and NHBA fused to the GNA1030 and GNA2091 proteins have been shown to enhance bactericidal activity compared to immunization with unfused proteins [28, 29]. While the functions of the *N. gonorrhoeae* homologues of GNA1030 and GNA 2091 are not characterized, these are also highly homologous to the recombinant proteins in 4CMenB.

4CMenB has been shown to induce antibodies that cross-react with *Neisseria gonorrhoeae* proteins, including outer membrane antigens, in both preclinical models and human sera. Mice vaccinated with 4CMenB had substantially accelerated *N. gonorrhoeae* clearance from the lower reproductive tract and reduced vaginal *N. gonorrhoeae* burdens compared to adjuvant only treated animals [30]. A follow-up study showed reproducible 4CMenB vaccine-mediated protection in a mouse model of upper genital tract colonization in which mice were challenged transcervically [31]. In humans vaccinated with 4CMenB, substantial IgG responses against gonococcal outer-membrane proteins (notably NHBA), and sera bactericidal activity against *N. gonorrhoeae* strains, have been demonstrated, confirming that vaccine-elicited antibodies cross-react with gonococcal antigens and contribute to anti-gonococcal killing *in* vitro [32, 33]. Altogether, these data support the biological plausibility of cross-species immunologic protection being induced by the vaccine. Since *N. gonorrhoeae* is an exclusive human pathogen, whose immunopathogenesis is not fully recapitulated by the mouse models, the next critical step is conducting human randomized controlled trials to formally determine whether 4CMenB prevents human gonococcal infection.

### Rationale for the use of human challenge

With this growing set of studies indicating that 4CMenB immunization provides protection from symptomatic *N. gonorrhoeae* infection, our group is evaluating the efficacy of 4CMenB against *N. gonorrhoeae* infection in a randomized controlled study under a Federal Drug Administration (FDA) Investigational New Drug (ClinicalTrials.gov Identifier: NCT05294588). We have a unique and innovative study design that incorporates the long-standing gonorrhea human challenge model, which will enable us to determine whether 4CMenB protects against *N. gonorrhoeae* infection or symptom development in the human male urethra.

The controlled human infection model (CHIM) of gonorrhea was developed concurrently at University of North Carolina at Chapel Hill (UNC-CH) and Walter Reed Army Institute of Research in the late 1980s and early 1990s [34]. It is safe and recapitulates the clinical features of naturally-acquired male gonococcal urethritis [35, 36]. Female participants are excluded from participating in studies that utilize the gonorrhea CHIM because of risk of ascending infection. The protocols utilized since the model’s inception have been subject to rigorous safety and ethical review by appropriate Institutional Review Boards (IRB (#21-0498, see *Ethical Justification* section below). Prior studies have demonstrated that wild-type *N. gonorrhoeae* strain FA1090, the challenge strain, has a dose-dependent capacity to cause productive urethral infection that is accompanied by signs and symptoms consistent with natural infection. Bacterial burdens and times to symptom development have all been well characterized. This model has also been used to test the importance of putative gonococcal virulence factors for urethral infection in men[35, 36]. Recently, our group conducted a randomized controlled trial testing the efficacy of a human monoclonal antibody that recognizes an abundant extracellular epitope present on *N. gonorrhoeae* and other bacteria in preventing experimental infection [37]. The *N. gonorrhoeae* CHIM has provided unique insights into infection dynamics, immunity, and treatment efficacy that cannot be obtained from observational studies or animal systems. We are now the only laboratory in the world currently using the gonorrhea CHIM platform.

### Ethical Justification

The intentional exposure of healthy adult volunteers to *Neisseria gonorrhoeae* raises important ethical considerations. Study protocols and procedures adhere to the ethical principles and regulations set forth in the Declaration of Helsinki and the U.S. Code of Federal Regulations for the Protection of Human Subjects. They also adhere to established frameworks on the acceptable use of controlled human infection studies, particularly the World Health Organization key criteria for ethical acceptability of CHIMs, including social/scientific value, risk minimization, informed consent, and independent review/oversight. The decision to conduct this study with the controlled human infection model is grounded in a rigorous risk-benefit analysis consistent with these frameworks and regulations and supported by more than three decades of safe implementation of the *N. gonorrhoeae* CHIM at the University of North Carolina at Chapel Hill (UNC-CH). Prior to this study, the UNC-CH controlled human infection model (CHIM) has been conducted safely in 257 male volunteers over 33 years without serious or irreversible adverse events. The challenge strain (FA1090) is fully characterized and susceptible to cephalosporin therapy.

The study ensures that the anticipated social and scientific values substantially outweigh the minimized and reversible risks to carefully selected, fully informed participants. Risks are minimized through established safety procedures, including rigorous eligibility screening, close daily clinical and microbiologic monitoring, 24-hour physician availability, and prompt, effective antibiotic treatment for *N. gonorrhoeae* with known susceptibility to the protocol directed therapy. These procedures, along with detailed inclusion/exclusion criteria and monitoring protocols, are described in the *Study Population* and *Description of Controlled Human Model* sections. Informed consent includes a comprehension assessment to ensure participants fully understand the nature of intentional infection and the associated risks and is obtained twice: first at enrollment and again immediately before inoculation. Independent oversight, interim safety review, and predefined withdrawal criteria further safeguard participants. Given the minimized, reversible risks and the high scientific and social value of this work, addressing an urgent public health need and generating critical vaccine efficacy and immune correlates data, the use of the CHIM is ethically acceptable.

### Added value of the CHIM approach

In parallel to the scientific and societal values, this study design offers unique advantages that make it an especially strong approach for evaluating *N. gonorrhoeae* vaccine efficacy at this stage of development that outperform over traditional Phase IIb study designs [37]. First, for clinical proof of vaccine efficacy, it addresses the limitations of animal models. *N. gonorrhoeae* is an obligate human pathogen, and no animal model fully reproduces the natural history, tissue tropism, or immune response seen in humans. The gonorrhea CHIM overcomes these constraints by enabling direct evaluation of vaccine-induced protection and immune responses in the natural host under controlled, reproducible conditions, albeit only in males.

Second, our design has the key advantages of offering a substantial increase in the power to detect protective efficacy. Because of exposure homogeneity, a dramatic reduction in the number of participants required to power a study to detect vaccine efficacy exists when compared to studying efficacy in the setting of naturally acquired infections (Table 1). Even for a partially effective vaccine, the sample size required for an adequately powered study to detect statistically significant effectiveness rises from less than 100 for a human challenge study to more than a 1000 in a very high-risk population and more than 25,000 in a population with average risk for *N. gonorrhoeae* infection (Table 1).

**Table 1.**
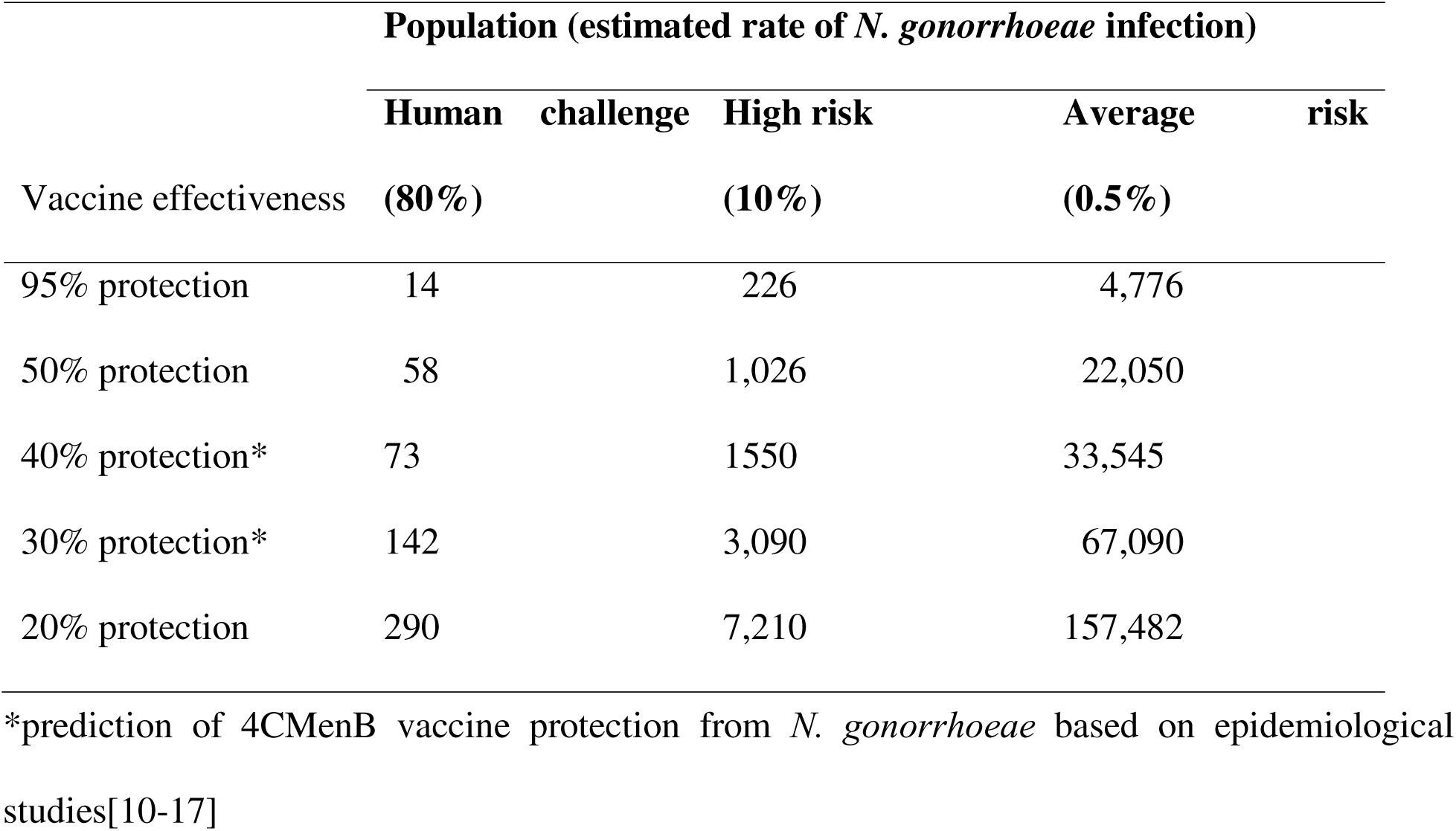
Estimated sample sizes for trials powered at 90% to detect vaccine efficacy for given levels of protection with one-sided alpha of 0.05 calculated using the Fisher Exact test for populations with 3 different proportions of *N. gonorrhoeae* infection during the trial.

Finally, our study also permits detailed investigation of early infection events and host immune responses under tightly defined conditions, providing a rare opportunity to identify immune correlates of protection in the natural human host. This trial will therefore not only deliver a direct efficacy estimate for 4CMenB against *N. gonorrhoeae* infection but also generate high-value mechanistic data to inform the design and prioritization of future vaccine candidates before embarking on large-scale field trials.

Building on these ethical and scientific advantages, we designed a randomized, double-blind controlled trial to directly assess the efficacy of 4CMenB against experimental *N. gonorrhoeae* infection. The following sections outline the study design, participant eligibility criteria, procedures, data collection, analysis, and dissemination, and safety oversight.

## Study design

The study is a single-site, double-blind randomized controlled trial testing efficacy of an FDA-approved 4-component MenB vaccine which contains *N. meningitidis* OMV (4CMenB) in preventing infection after intraurethral challenge with *N. gonorrhoeae* (Figure 1). We hypothesize that two doses of the FDA-approved 4CMenB given intramuscularly at least 1 month apart protects from intraurethral challenge with *N. gonorrhoeae* in men. The control arm of the study provides immunization with two commercially available FDA-approved vaccines that do not have relevance to *N. gonorrhoeae* infection: quadrivalent influenza (Flu) vaccine (FLULAVAL^TM^) and tetanus/diphtheria (Td) vaccine (TDVAX^TM^). The effect of the intervention will be compared to two doses of FDA-approved control vaccines, namely one dose of the Flu vaccine and one dose of the Td vaccine. The objectives and outcome measures are detailed in Table 2.

**Figure 1.**
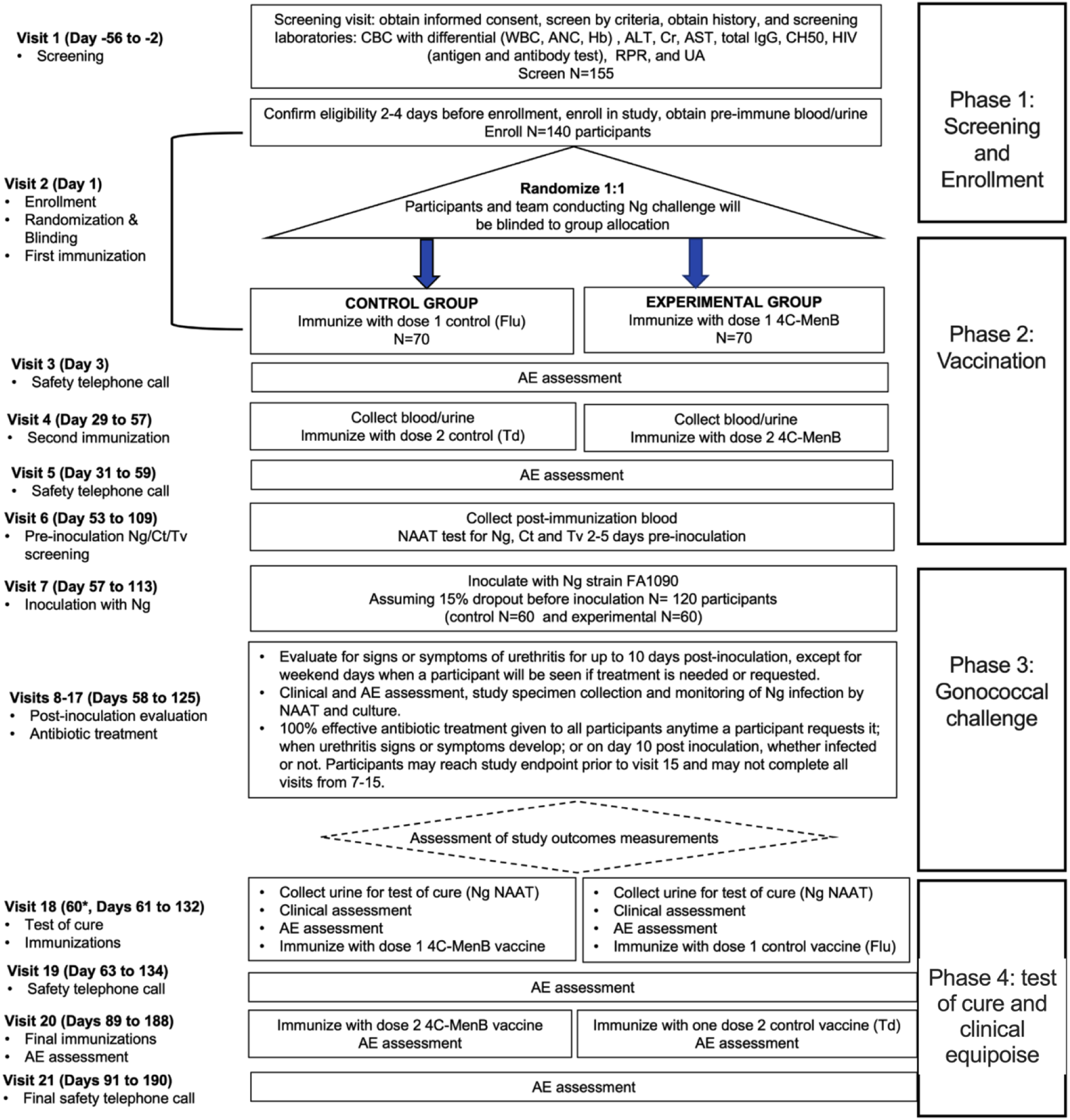
Interventions, study visits, and participant timeline.

**Table 2.** Objectives and outcome measures.

| Objective | Outcome | Outcome Measure |
| --- | --- | --- |
| **Primary** |  |  |
| To determine the efficacy of standard immunization with 4CMenB in preventing experimental urethral infection with *N. gonorrhoeae* in men | *N. gonorrhoeae* infection in each study group<br>**Definitions:**<br>Infection is defined by positive *N. gonorrhoeae* NAAT or positive urine culture or positive swab culture on the day of antibiotic administration.<br>A participant is deemed protected when not infected with *N. gonorrhoeae*, as defined by negative *N. gonorrhoeae* NAAT and negative urine culture and negative swab culture on the day of antibiotic administration.<br>Evaluable antibiotic administration days can occur anytime 1-10 days post-urethral inoculation with *N. gonorrhoeae*.<br>If a participant requests antibiotic treatment on the same day as urethral inoculation, he will not be evaluable for primary or secondary study outcomes but may be evaluable for exploratory outcomes of immune responses to vaccination. | Infectivity at the time of antibiotic treatment for urethritis, or 10 days post-urethral inoculation with *N. gonorrhoeae* if signs or symptoms do not develop, in each of the two study groups.<br>**Definitions:**<br>Infectivity is defined as the proportion of participants positive for *N. gonorrhoeae* on antibiotic treatment day by *N. gonorrhoeae* NAAT or urine culture or swab culture in each study group. |
| **Secondary** |  |  |
| To determine the effect of 4CMenB immunization on the development of urethritis caused by experimental *N. gonorrhoeae* infection<br>To determine the effect of 4CMenB immunization on the time to onset of urethritis caused by experimental *N. gonorrhoeae* infection | Symptomatic *N. gonorrhoeae* infection in each study group<br>**Definitions:**<br>Symptomatic *N. gonorrhoeae* infections are defined in two ways:<br>Macroscopic urethritis, defined as clinician-observed urethral discharge on day of antibiotic treatment.<br>Symptomatic microscopic urethritis, defined as the presence of dysuria or participant-observed urethral discharge or urethral discomfort accompanied by $\geq 5.8 \text{ Log}_{10} \text{ WBC/mL}$ urine sediment (equivalent to $\geq 4 \text{ WBC/high power field}$) on day of antibiotic treatment.<br>Macroscopic and symptomatic microscopic urethritis are assessed daily for up to 10 days after experimental *N. gonorrhoeae* inoculation.<br>An asymptomatic infection is defined by a positive *N. gonorrhoeae* NAAT, positive urine culture, or a positive swab culture on antibiotic treatment day in the absence of *N. gonorrhoeae* symptoms. | Proportion of patients who develop of macroscopic urethritis<br>Proportion of patients who develop symptomatic microscopic urethritis<br>Time to onset of macroscopic urethritis (number of days after urethral inoculation that antibiotic treatment is triggered by macroscopic urethritis) |

The study consists of 4 phases: screening and enrollment, immunization, *N. gonorrhoeae* challenge and daily post-inoculation evaluations for infection, and follow-up immunization. Each of these phases are explained in detail in the study protocol (Supplementary Information). Individual participation in all four phases requires at least approximately 4 months. In the final immunization phase, all participants are crossed over to receive the immunizations they did not receive in the first immunization phase. The participants in the control arm receive two doses of 4CMenB according to label instructions at the time of study design to achieve clinical equipoise. The participants in the experimental arm receive one dose of Flu vaccine and one dose of Td vaccine according to label instructions.

## Study Participants

Men aged ≥18 and <36 years who have not previously been received a MenB vaccine will be recruited from local populations in Chapel Hill, Durham, and surrounding areas. Due to potential complications from ascending gonococcal infection following bacterial challenge, women were excluded from the study. Because the feasibility of this proposed study is dependent on a population lacking a history of 4CMenB vaccination, eligible participants who have previously received 4CMenB immunization are also excluded from this study. Participants with no medical contraindication to 4CMenB administration, no contraindication to intraurethral challenge with *N. gonorrhoeae*, and who are willing and able to consent and participate in the study will be enrolled in this study (UNC IRB #21-0498).

Participants in this protocol provide written informed consent twice: once with an accompanying comprehension test at screening before any study procedures are performed and a second time on the day of inoculation just prior to inoculation. Inclusion and exclusion criteria (section 5.1 of Study Protocol in Supplementary Information) is confirmed by the study physician or designee licensed to make medical diagnoses. To ensure participant safety, individuals are excluded if they face any significant risk from vaccine receipt or inoculation with *N. gonorrhoeae*. For example, all participants are screened for complement deficiency, a known risk factor for disseminated gonorrhea. Those taking medications with known interactions with antibiotics used for gonorrhea treatment, using immunomodulators that could alter vaccine responses or infection risk, or with urethral abnormalities preventing safe catheter insertion were excluded. In addition, any clinical adverse event, laboratory abnormality, intercurrent illness, or other medical condition arising after enrollment that could compromise participant safety or study integrity warranted removal from the study.

First study screening was conducted on April 24, 2022. Data collection will conclude by the end of January 2026. Final safety follow-up procedures and vaccinations for equipoise will conclude in spring 2026.

## Vaccine and antibiotic sourcing

All study-specific vaccines and antibiotics are procured, stored, handled, and dispensed by the UNC Investigational Drug Services (IDS) in accordance with institutional procedures and ICH-GCP guidelines. Documentation of receipt, inventory, and accountability is maintained in accordance with GCP standards. IDS utilizes the UNC Pharmacy Support Services (PSS) as its procurement intermediary. PSS sources medications primarily from AmeriSource Bergen, the principal supplier for all UNC Health pharmacies. Medications are typically delivered from PSS to IDS within 48 hours of order placement, ensuring timely availability for study visits and interventions. For the seasonal influenza vaccine (Flulaval), IDS coordinates annually with the Assistant Director of the UNC Health Care System Pharmacy Supply Chain. In the spring, IDS submits a request for priority allocation of the upcoming year’s formulation. Upon regulatory approval in late summer, GlaxoSmithKline (GSK) provides an initial supply of Flulaval directly to IDS.

## Study procedures

### Master bank of bacterial product

*N. gonorrhoeae* strain FA1090 has been used extensively in experimental infection studies conducted at UNC [34–36, 38–41]. FA1090 is resistant to streptomycin (Sm^R^) and sensitive to ciprofloxacin and cefixime. *N. gonorrhoeae* strain FA1090 variant A26 is being used for the current study. A master bank of this variant has been generated by preparing a bulk culture under controlled conditions, cryoprotected, dispensed into labeled cryovials, and stored at ≤–80 °C with inventory control. It was validated by lot-release testing, conducted according to Good Laboratory Practices as described in the Investigator’s Brochure and Chemistry, Manufacturing and Controls documents. Lot-release testing (identity, purity/sterility, viability/titer, antimicrobial susceptibility profiling, and other required phenotypic markers) was completed per predefined acceptance criteria prior to release for use in inocula preparation. The *N. gonorrhoeae* to be used for this clinical trial has been reviewed and approved by the UNC Institutional Biosafety Committee and by the FDA under a sponsor-investigator IND.

### Inoculum preparation

The procedures governing the inoculum preparation have been previously described [40]. In brief, we thaw a lot-released single-use viable of cryopreserved *N. gonorrhoeae* strain FA1090 variant A26 and grow bacteria overnight on solid medium. The next day (day of inoculation), we use colony phenotypic characteristics to select predominantly piliated, Opacity protein (Opa)-negative gonococci expressing the lacto-*N*-neotetraose LOS epitope recognized by monoclonal antibody (Mab) 3F11, which is associated with development of urethral discharge in natural and experimental gonococcal infection in men[42]. We prepare a liquid suspension of bacteria and inoculate study participants with an target delivered dose of approximately 1.0 x 10^6^ colony forming units [log10(6)± 1 log□□]. This dose maximizes the number of evaluated infections while reducing the risk of inducing urethritis due to microbial load without establishing infection. Based on prior knowledge and established dose response curves, this dose is expected to lead to infection of ∼80% of the inoculated participants (ID80) [34, 40].

### Blinding

At enrollment, participants are randomized in a 1:1 ratio to the control vaccine arm or the experimental vaccine arm. To ensure balanced enrollment between trial arms, randomization will occur in blocks of 4. Group assignment is made according to a computer-generated random sequence by the study statistician. At the time of study initiation, the study statistician created a blinded version of the block randomization list that only lists the random codes (allocation assignment column removed) at time of study initiation. When enrolling a participant, the blinded clinical staff assigns the participant the next sequential code from the blinded randomization list. They document the assignment of this code on their blinded randomization list. The blinded clinical staff communicates the assigned code to the unblinded pharmacy staff. The unblinded pharmacy staff documents the assignment of this code on their unblinded randomization list, referencing the allocation assignment that corresponds with the code. They prepare vaccination for participant according to this allocation assignment and provide the prepared vaccines in a stapled brown paper bag. The bag is given to the blinded staff who is administering the vaccines. The unblinded staff do not open the bag or see the vials. Emergency unblinding by the site Investigator of Record (IoR) is only undertaken if the information is needed for immediate medical management of the participant. A participant is unblinded only if knowing the assigned treatment is critical for making immediate therapeutic decisions. If emergency unblinding is indicated, the IoR must consult study team and DSMB in advance of the unblinding. The IoR informs the Pharmacist of Record (PoR) of the requested unblinding in writing. The PoR provides the participant’s allocation assignment to the IoR for immediate medical management of the participant. The IoR only communicates the unblinded vaccination information to the participant if this is indicated.

### Vaccination procedures

Vaccines are administered in accordance with label instructions and the CDC recommendations that were active at the time of study initiation, which for 4CMenB was prior to the updated vaccine schedule that lengthened the time between the two doses of 4CMenB.

### Inoculation and post-inoculation procedures

After the first immunization phase is completed, participants are challenged with *N. gonorrhoeae* within a 2-10 week window and monitored for development of *N. gonorrhoeae* infection for 10 days. Because we are monitoring the participants for up to 10 days post-challenge for *N. gonorrhoeae* infection, irrespective of symptoms, the human model of urethral infection can assess both microbiological outcomes (i.e., do participants acquire infection with recoverable viable bacteria?) and disease outcomes (i.e., do participants develop urethritis after inoculation?). In doing so, we will be able to rule out whether the 4CMenB vaccine is associated with asymptomatic infection or delayed development of infection rather than protection. Among all experimentally infected participants prior to the current study, all individuals inoculated with wild type *N. gonorrhoeae* (strain FA1090) that developed infection also experienced evidence of clinical urethritis within 5 days after bacterial inoculation.

Prior to inoculation the procedure is reviewed with participants and informed consent is obtained a second time to reaffirm the subject’s continued willingness to participate in the challenge phase of the study. Inoculation is accomplished by instillation of ∼0.3 mL of a *N. gonorrhoeae* suspension through a pediatric feeding catheter (8fr) inserted ∼5 cm into the urethra. The inoculation procedures have been previously described [40]. For this study we have developed and deployed an outpatient protocol in which participants are inoculated, discharged, and instructed to return for observation daily, thus increasing clinical research capacity as opposed to an inpatient protocol. Infection is monitored by nucleic acid amplification-based (NAAT) testing, using an FDA-approved clinical test, and bacterial culture from daily urine specimens. Daily urine dipstick analysis and microscopic examination of urine sediment for white blood cells / polymorphonuclear cells as an indicator of urethral inflammation will also be performed. Before antibiotic treatment on Day 10 post-inoculation, or the day on which a participant is treated during the trial, a urethral swab in addition to the second urine specimen urine will be collected. Clinical disease is monitored by daily targeted genital exam. Symptomatic infection or *N. gonorrhoeae* urethritis will be defined as the development of urethral discharge observed by the study physician or the report of urethral symptoms (discharge or dysuria) reported by the patient in conjunction with pyuria≥5.8 log_10_ WBC/mL urine sediment on the day of antibiotic treatment. Antibiotics used in this study (ceftriaxone or cefixime) were selected based on demonstrated full susceptibility of the *Neisseria gonorrhoeae* strain used in the inoculum. No treatment failures with ceftriaxone or cefixime have been observed in prior human challenge studies. All participants receive prompt antibiotic treatment with a third-generation cephalosporin (single 400 mg oral dose of cefixime or single 250 mg dose delivered intramuscularly of ceftriaxone) after challenge with *N. gonorrhoeae* when: (1) requested by an individual participant, regardless of signs, symptoms or positive cultures; (2) purulent urethral discharge is observed by the examining clinician or reported by the participant; or (3) when the challenge period is complete (on day 10 after inoculation), whether infected or not. Antibiotic treatment marks the end of the challenge/post-inoculation evaluation phase A test of cure is performed within 7 days of antibiotic treatment. From previous *N. gonorrhoeae* challenge experiments, three days after antibiotic treatment is the mean time to test of cure administration.

### Adverse events and withdrawals

Participants are monitored for AEs during the entire study duration (from study enrollment until study completion). All AEs that occur from vaccination and/or inoculation through the final follow-up visit will be reported. A study physician is always on call during experimental infection trials. If participants are withdrawn prior to challenge or after challenge but before test of cure, they will not be evaluable for study outcomes. If they are withdrawn after test of cure, they will be included in outcome evaluation.

### Sample size

For this trial, we estimate that the primary outcome (infection) will occur in 80% of the control group, based on previously determined infectious dose (ID_80_) of the inoculating *N. gonorrhoeae* strain FA1090 and the inoculum size to be used for the challenge (10^6^ CFU) [34, 40]. Previous retention in experimental infection studies has been high (1 requested withdrawal of 256 enrolled participants in the previous 27 years). To account for extended study participation, potential vaccine-related reactogenicity, and the possibility of dropout prior to challenge, we assume a 15% attrition rate between enrollment and challenge. Accordingly, we plan to enroll 140 participants to achieve 120 evaluable participants (60 per arm). With this design, we have 80% power at a one-sided 5% type I error to detect 30% vaccine efficacy, calculated as:

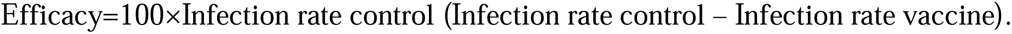

The intention-to-treat principle will be applied. If infection occurs in only 70% of controls, power falls to ∼70%; if infection occurs in 90% of controls, power rises to 96% to detect 30% efficacy. Based on prior study data, the likelihood of developing urethritis when infection occurs is 100%. Because urethritis develops in nearly all infections, power to detect a reduction in urethritis will be similar to that for infection. However, secondary analysis of time to urethritis will be underpowered, since we expect ≥20% of participants not to experience the event, making time-to-event comparisons less reliable.

### Statistical Analysis Plan

We will first summarize all major variables and check data quality, including distributions and missingness. If missing data are minimal and random, we will analyze available cases; multiple imputation will be used as a sensitivity check. Categorical variables may be grouped, and continuous variables may be transformed or analyzed with non-parametric methods as needed. To confirm randomization, baseline measures will be compared between study arms. Any imbalances with potential to bias results will be adjusted for, though we anticipate little confounding. Analyses will follow the intention-to-treat principle, accounting for ∼15% dropout. Results will be reported with confidence intervals; p<0.05 will be considered statistically significant, while higher p-values will be reported as inconclusive. For the primary endpoint (infection), we will compare infection rates between vaccine and control groups using a one-sided Fisher exact test. Secondary endpoints (urethritis symptoms) will be analyzed in the same way. Time to macroscopic urethritis will be visualized and compared between groups using survival analysis (log-rank test).

### Data collection and management, and confidentiality

The site principal investigator and delegated study personnel are responsible for collecting and documenting all protocol-required data points. Study personnel record clinical study data on site-controlled source documentation forms. The source documentation forms are designed by study personnel according to the protocol to ensure all required assessments are correctly performed and appropriately documented. Source documentation is completed per Good Documentation Practices (GDP) principles and ALCOA+ guidelines. Study personnel create a research record for every participant who signs consent. Completed source documents are maintained in the participant’s research record. The principal investigator performs routine reviews of all study data for each participant and signs off in the research record to document this review. Site data staff perform quality control reviews of all source documentation and lab reports. After verifying the documents for completeness and accuracy, data staff enter the data into the study database’s electronic Case Report Forms (eCRFs). The database is a validated, 21 Code of Federal Regulations (CFR) Part 11 compliant electronic data capture system, which is maintained by site data staff. The database includes password protection, audit trails, and internal quality checks to identify data that appear inconsistent, incomplete, or inaccurate. After the initial written informed consent is provided, site personnel assign a unique study code number to each participant to ensure confidentiality. This code is used to ensure de-identification of study data recorded on the participant’s source documents, research specimens, test results, and eCRFs. The log that links the unique study codes to identifying participant information is maintained on a password-protected computer file that is only accessible by authorized personnel directly involved with the study. Only authorized personnel directly involved with the study will have access to individually identifiable information about participants. All participant research records are stored in locked cabinets within the site’s administrative office, which is only assessable to designated study personnel via key card. The filing cabinets are unlocked only when in use by the study personnel and remain locked at all other times.

### Quality Assurance and Protocol Deviations

The principal investigator will review study records regularly, and data staff will perform quality checks on data entry. Protocol deviations will be documented, reported to the IRB as required, and corrective actions implemented. Data and Safety Monitoring Board (DSMB). Safety oversight will be under the direction of a Data and Safety Monitoring Board (DSMB), as detailed in the study protocol (Supplementary Information).

### External monitoring

Monitoring will follow a written clinical monitoring plan. Site initiation and interim visits will include review of regulatory files, participant records, and laboratory data to ensure protocol adherence and data integrity.

### Data Availability Plan

In recognition that data sharing is essential for expedited translation of research results into knowledge, products, and procedures to improve human health, we intend to share final research data with the wider scientific community. De-identified research, including a data dictionary, will be made available when the study is completed and published in a public repository such as UNC Dataverse. Summary results will also be made available on ClinicalTrials.gov.

### Patient and public involvement

Patients or members of the public were not involved in the design, conduct, or reporting of this controlled human infection trial. This study targets a very specific population of healthy adult male volunteers without prior MenB vaccination, and the protocol was instead developed by an experienced multidisciplinary clinical and methodological team and reviewed by the institutional review board and Data and Safety Monitoring Board.

## Discussion

This study represents an important advancement in *Neisseria gonorrhoeae* vaccinology by directly testing whether 4CMenB, a licensed *Neisseria meningitidis* serogroup B OMV-based vaccine, can prevent gonococcal infection or urethritis. The unique study design leverages our group’s long-standing controlled human infection model (CHIM) of *N. gonorrhoeae* urethral infection, through which vaccine efficacy assessment can be streamlined under optimized and rigorously defined infection conditions. This offers multiple benefits over traditional, costlier Phase IIb trials.

### Strengths and limitations

A principal advantage of the CHIM is statistical efficiency. Retrospective population-based studies have indicated that mass immunization with OMV-based MenB vaccines, including 4CMenB, was associated with a 30-40% reduction in symptomatic gonorrhea rates. For vaccines with partial efficacy, like 4CMenB, the required sample size to demonstrate protection is substantially reduced in the challenge setting compared with natural exposure studies. In addition, controlled exposure ensures more uniform timing of vaccination, infection, and follow-up, minimizing confounding from waning immunity or variable risk behaviors. However, the data generated by these retrospective, observational studies cannot disentangle whether the observed effect reflects the vaccine’s ability to prevent infection or simply an attenuation of clinical disease, since these studies did not prospectively test for *N. gonorrhoeae* and have relied on clinic-based data. The present study is designed to address this uncertainty. While the primary objective is to determine whether 4CMenB prevents acquisition of infection (defined as infection detected by *N. gonorrhoeae* nucleic acids in self-collected urine and/or recovery of live *N. gonorrhoeae* from culture), the secondary analyses will clarify whether vaccine-induced effects are limited to delaying or reducing symptomatic urethritis.

Historically, participants in our intraurethral challenge developed clinical urethritis by day 3 post-inoculation, on average. This is consistent with the natural clinical course of infection in males, who develop symptoms 2-7 days after exposure [43–45]. Daily follow-up up to 10 days post-inoculation, including the collection of two independent urine specimens daily, enable a comprehensive assessment of infection acquisition and clinical presentation outcomes.

An additional strength of the design is the ability to investigate immune correlates of protection in the natural host. Serum bactericidal activity is the established surrogate of protection for invasive serogroup B meningococcal diseases but may not reflect protection at the genital mucosal sites where *N. gonorrhoeae* primarily colonizes. The true correlate of protection of 4CmenB against serogroup B meningitis remains unclear. 4CMenB does induce gonococci-reactive antibodies targeting key vaccine antigens in mice and humans, but the exact correlates of protection are still lacking. Collection of blood and urine at defined intervals relative to vaccination and challenge will permit exploratory analyses of systemic and local immune responses, supporting the identification of correlates of protection relevant to the male urethra.

Several limitations must be acknowledged, all of which primarily affect the generalizability of the findings, with respect to strain, anatomical site of infection (urethral vs. cervicovaginal, genital vs. extragenital) and vaccination regimen. First, the study assesses efficacy against a single strain (FA1090). Although antigenic variation remains a critical barrier to vaccine development, OMV-based vaccines present diverse antigens, including N*. gonorrhoeae* homologs of recombinant MenB components, which may broaden protective responses. Second, the protocol was initiated under a schedule of two doses given 28 days apart. In August 2024, the Food and Drug Administration (FDA) changed the label for 4CMenB from the 2-dose schedule to a 2-dose scheduled with a longer (6 month rather than 1 month) interval between doses or a 3-dose schedule (0, 1–2, and 6 months), based on new immunogenicity data. Consequently, the findings of this study may only apply to individuals who underwent vaccination prior to August 2024. Third, participation is restricted to men due to ethical considerations, excluding evaluation in females despite the greater long-term health consequences of infection in women. Fourth, the study focuses exclusively on urethral infection and does not address extragenital sites, such as pharyngeal or rectal infection, which are critical reservoirs for transmission and may respond differently to vaccination.

Currently, this study relies on enrolling individuals without prior MenB immunization. However, if a *N. meningitidis* B outbreak occurs, it may be unethical to withhold 4CMenB from at-risk populations. In such a scenario, we would consider incorporating the MenB-fHbp vaccine as a control, given that it lacks cross-reactive *N. gonorrhoeae* surface antigens, allowing for evaluation of the specific effects of 4CMenB. If widespread MenB vaccination becomes standard, alternative study designs may be required, such as evaluating the efficacy of a booster dose in previously vaccinated individuals compared to a Tetanus-Diphtheria booster.

## Conclusion

Vaccine development against *N. gonorrhoeae* has been particularly challenging in the past due to the pathogen’s immune evasion strategies and the absence of durable natural immunity in humans. Through use of the long-standing, and safe controlled human challenge model that recapitulates the natural course of *N. gonorrhoeae* urethral infection, we will directly test whether, under defined conditions, an already FDA-approved vaccine containing *N. meningitidis* OMV prevents infection, delays onset, or reduces symptomatic disease. By combining precise infection timing, intensive clinical and microbiological monitoring, and systematic immune sampling, the resulting well-characterized cohort and accompanying immunological data will provide critical insights into correlates of protection and inform the development of next-generation vaccines against *N. gonorrhoeae*, whether OMV-based or derived from alternative platforms.

## Supporting information

SPIRIT checklist

## Acknowledgments

We thank the study participants for their time and commitment. We acknowledge the University of North Carolina at Chapel Hill Clinical Trials Unit (CTU) and Clinical and Translational Research Center (CTRC) for their essential support in study implementation, as well as the UNC Center for AIDS Research (CFAR) for scientific and infrastructure support.

